# COVID-19 critical care simulations: An international cross-sectional survey

**DOI:** 10.1101/2020.11.17.20233262

**Authors:** Mohamad-Hani Temsah, Abdulkarim Alrabiaah, Ayman Al-Eyadhy, Fahad Al-Sohime, Abdullah Al Huzaimi, Nurah Alamro, Khalid Alhasan, Vaibhavi Upadhye, Amr Jamal, Fadi Aljamaan, Yaseen M Arabi, Marc Lazarovici, Abdulaziz M. Boker

**Author notes:** These authors contributed equally to this work (Mohamad-Hani Temsah, Abdulkarim Alrabiaah). **Corresponding author:** Amr Jamal, College of Medicine, King Saud University, King Saud University Medical City, PO Box 2925, Riyadh 11461, Saudi Arabia.

## Abstract

**Introduction:** Many healthcare systems initiated rapid training with COVID-19 simulations for their healthcare workers (HCWs) to build surge capacity and optimize infection control measures. This study aimed to describe COVID-19 simulation drills in international healthcare centers.

**Methods:** This is cross-sectional, international survey among simulation team leaders and HCWs, based on each center’s debriefing reports from simulation centers from 30 countries in all WHO regions where COVID-19 simulation drills were conducted. The main outcome measures were the COVID-19 simulations characteristics, outcomes, facilitators, obstacles and challenges encountered during the simulation sessions.

**Results:** Invitation was sent to 500 simulation team leaders and HCWs, 343 responded, and 121 completed the survey. Those who completed the study were from East Mediterranean (EMRO) countries (41.3%); Southeast Asian countries (SERO) (25.6%); and Europe (12.4%) and the remainder from other regions. The frequency of simulation sessions was monthly (27.1%), weekly (24.8%), twice weekly (19.8%), or daily (21.5%). Among participants, 55.6% reported team’s full engagement in the simulation sessions. The average session length was 30–60 minutes. The most commonly reported debriefing leaders were ICU staff, simulation lab staff, and ER facilitators, and the least common were infection control staff. A total of 80% reported “a lot” to “a great improvement” in terms of clinical preparedness after simulation sessions, and 70% were satisfied with the COVID-19 simulation sessions and thought they were better than expected. Most of the perceived issues reported were related to infection control measures, followed by team dynamics, logistics, and patient transport issues.

**Conclusion:** Simulation centers team leaders and HCWs reported positive feedback on COVID- 19 simulation sessions. The presence of multiprofessional personnel during drills is warranted. These drills are a valuable tool for rehearsing safe dynamics of HCWs on the frontline of COVID-19.

**Summary box:** explaining the significance of their study by providing each of the following key questions:

**What is already known?:**

- Simulation enhances healthcare systems safety.
- Preparedness to potential disasters includes training for personal protection techniques, environmental contamination, medical management, and training of HCWs.

**What are the new findings?:**

- Many hospitals conducted COVID-19 simulations in all WHO regions.
- Most of the team leaders and HCWs reported full engagement and significant clinical preparedness improvement after the COVID-19 simulation sessions.

**What do the new findings imply?:**

- The presence of multiprofessional personnel, including infection control experts, during COVID-19 drills is warranted.
- Simulation are a valuable tool for rehearsing safe dynamics of HCWs on the frontline of COVID-19.

## Introduction

When the COVID-19 crisis started in early January 2020, the World Health Organization (WHO) reported cluster of pneumonia cases in Wuhan, China [(Organization, 2020)]. Soon it has increased exponentially till the disease was declared a pandemic by the WHO in March which urged healthcare systems to initiate rapid training for their healthcare workers (HCWs) to cope with this rapidly evolving pandemic.

Preparedness to such potential disasters includes training for personal protection techniques, environmental contamination, medical management, training of recruited staff, ethical issues, and psychosocial issues as recommended by The European Society of Intensive Care Medicine’s Task Force for Intensive Care Unit [(Richards and Sprung, 2010)]. All these aspects in addition to debriefing reports could be assets to healthcare administration and policymakers. A Cochrane review concluded that face-to-face training in PPE use might reduce errors better than video- or folder-based training [(Verbeek et al., 2019)].

During the 2003 severe acute respiratory syndrome (SARS) crisis, Abrahamson et al. proposed and tested a novel protocol for cardiac arrest in a patient with SARS. The protocol was promptly and effectively instituted by teamwork training in high-fidelity simulation drills [(Abrahamson et al., 2006)]. Previous models that addressed intensive care units and hospital preparations for an influenza epidemic or mass disaster suggested enforcing communication, coordination, and collaboration between the ICU and key interface departments [(Sprung et al., 2010)].

Simulation is an important training tool that could especially be used to test and boost preparedness to global pandemics or natural disasters or man-made mass casualties incidents [(King et al., 2006)]. Therefore, sharing the most common debriefing findings from such simulations could benefit other sites that did not have such simulations. During the pandemic, rapid sharing of global experiences could help other healthcare providers and policymakers expedite solutions to rectify commonly encountered errors. This could improve patients’ outcomes and improve infection control measures to prevent such errors from occurring in centers where no COVID-19 simulation drills were performed [(Aldekhyl and Arabi, 2020)].

COVID 19 has been labeled by the WHO as a novel infection that spread quickly across the globe and is associated with high fatality and transmission rates, its unclear characteristics regarding transmission and infection control precautions initially had been alarming and mandated simulation programs of different settings and high preparedness standards in order to face its high spread rate.

There is limited data on the use of simulation in the response to COVID-19 pandemic around the world. Therefore, we conducted this international survey to describe the characteristics of COVID-19 simulation sessions (drills) among international healthcare centers where COVID-19 simulations were conducted, explore the facilitators and barriers to COVID-19 simulations, and explore participants’ feedback on COVID-19 simulations.

## Method

Data collection was conducted using an online survey on Survey Monkey ®. The questionnaire was developed by the research team following multidisciplinary team meeting and brain storming of simulation experts for relevant questions to add into the survey based on their expertise in the medical simulation setting and on the debriefing reports’ from the three simulation centers. The content validity of the questionnaire was tested with another four simulation lab experts.

### Site recruitment

Hospitals equipped with simulation centers across 30 countries of the WHO regions were contacted wherever COVID-19 simulation drills were performed [(Alleva et al., 2020)]. Participants were invited either by direct contact of international simulation center leaders or via emails to simulation center societies (listed at https://www.ssih.org/Home/SIMCenter-Directory).

### Study population

The inclusion criteria comprised simulation team leaders and HCWs who participated in drills that were performed specifically for COVID-19 case handling that were followed by debriefing in the recruited sites. The HCWs include physicians, nurses, respiratory therapist or technologists who attended the meeting. Simulation sites or participants without documented debriefings were excluded.

### Enrollment and consent

Before participation, the purpose of the study was explained in English at the beginning of the electronic survey. The respondent was given the opportunity to ask questions via a dedicated email address for the study. The study was approved by the institutional review board at King Saud University approved the study. Waiver for signed consent were obtained, since the evaluation presented no more than minimal risk to subjects and involved no procedures for which written consent is usually required outside the study context. To maximize confidentiality, personal identifiers were not required.

Data collection was conducted using the survey items. Following an extensive literature review of previous simulation studies’ questionnaires, the survey items were adopted and modified to answer the current research questions [3, 4]. The participants were asked about characteristics of COVID-19 simulation drills at their site, facilitators and barriers to COVID-19 simulations, and feedback on the COVID-19 simulations.

### Patient and Public Involvement

No patients were involved in this research.

### Data management and analysis

Data collection was completed based on electronic surveys utilizing the questionnaire. The data from the questionnaires were transferred into an Excel database. The data was then cleaned and analyzed using SPSS.

### Statistical Data Analysis

Means and standard deviations were used to describe the continuous variables and the frequencies and percentages for the categorically measured variables. A multiple response dichotomy analysis was used to describe the questions that allowed selection of more than an option. Statistical analysis was done using Statistical Package for the Social Sciences (SPSS) version 21 for windows 8.1. Microsoft Excel V16.43.1® was used for the creation of figures and depictions.

## Results

From the 500 participants, 343 responded. However, only 121 participants completed the survey and were included in the analysis. The majority of respondents were female (62.8%), and their age distribution was mainly 35–44 years and 45–54 years, with these groups accounting for 29.8% and 33.1%, respectively. (Table 1)

**Table 1:** Descriptive analysis of the healthcare workers’ sociodemographic and professional characteristics. N = 121.

|  | Frequency | Percentage |
| --- | --- | --- |
| **Sex** |  |  |
| Female | 76 | 62.8 |
| Male | 45 | 37.2 |
| **Age (years)** | 121 |  |
| 18–24 years | 16 | 13.2 |
| 25–34 years | 29 | 24 |
| 35–44 years | 36 | 29.8 |
| 45–54 years | 40 | 33.1 |
| **Clinical Role** |  |  |
| Physician | 68 | 56.2 |
| Nurse and Respiratory therapist | 53 | 43.8 |
| **Country of practice** |  |  |
| Europe (EURO) Region | 15 | 12.4 |
| East Mediterranean (EMRO) Region | 50 | 41.3 |
| Africa (AFRO) Region | 7 | 5.8 |
| Region of the Americas (PAHO) | 8 | 6.6 |
| South-East Asia (SERO) Region | 31 | 25.6 |
| Western Pacific Region (WPRO) Region | 10 | 8.3 |
| **Role in the simulation experience:** |  |  |
| Simulation Team Leader/Organizer | 29 | 24 |
| Healthcare provider attending/participating in the simulation | 92 | 76 |

### Demographic of the respondents

More than half of the respondents (56.2%) were physicians. Most (76%) of the respondents were HCWs who were involved in simulation drills related to COVID 19 while only 24% were simulation teams’ leaders/organizers. The distribution of the areas of practice of the respondents was: 41.3% from the East Mediterranean (EMRO) countries region, 25.6% from the Southeast Asian countries (SERO) region, 12.4% from Europe, and the remainder from other regions (Figure 1) [(WHO)].

**Figure 1-A:**
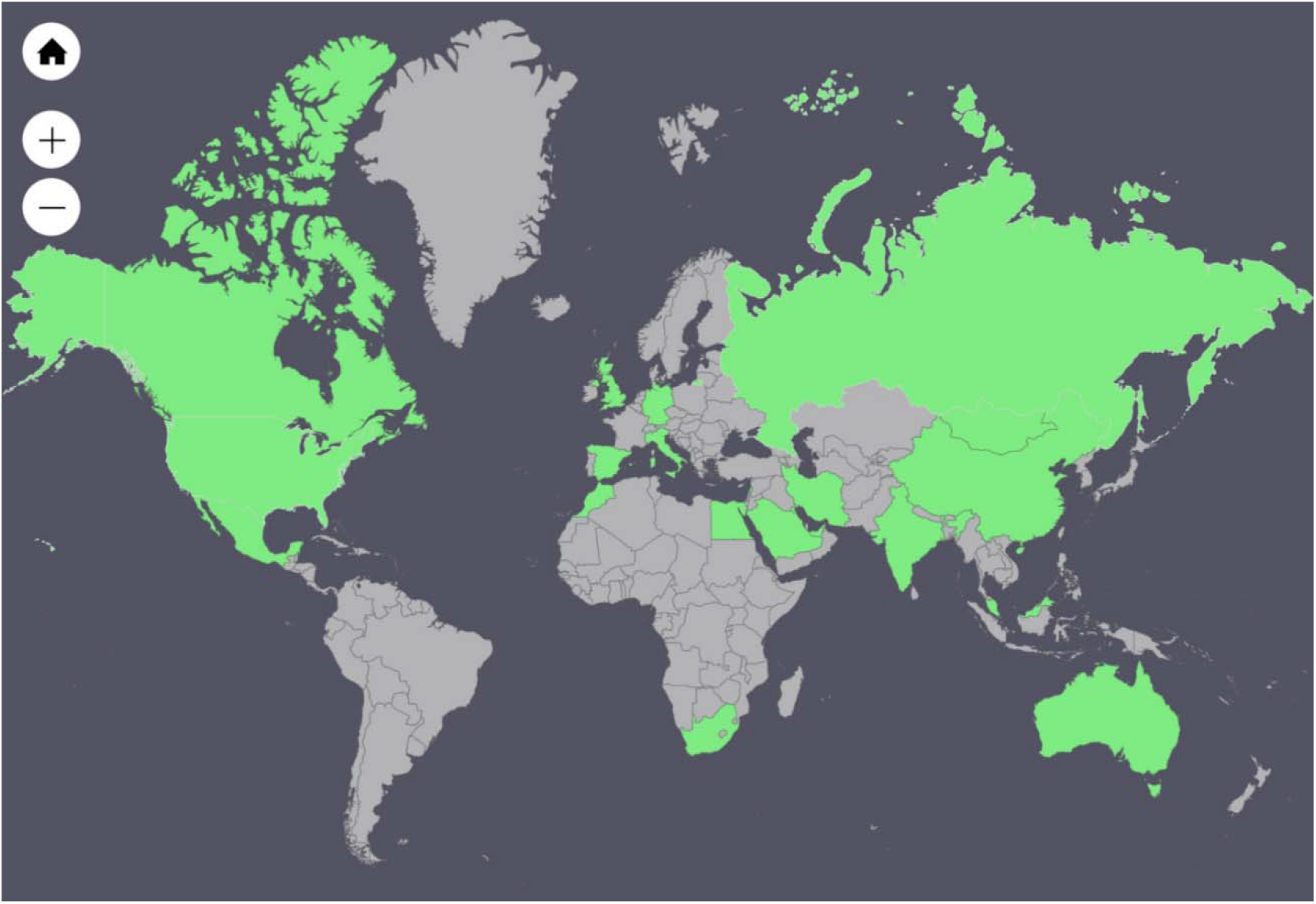
Respondents’ reports of the countries where their COVID-19 simulation was conducted*. * Afghanistan, Australia, Barbados, Canada, China, Egypt, Germany, India, Italy, Kuwait, Lebanon, Malaysia, Mexico, Mongolia, Morocco, Russian Federation, Saudi Arabia, Singapore, South Africa, Spain, United Arab Emirates, United Kingdom of Great Britain and Northern Ireland, United States of America

**Figure-1B:**
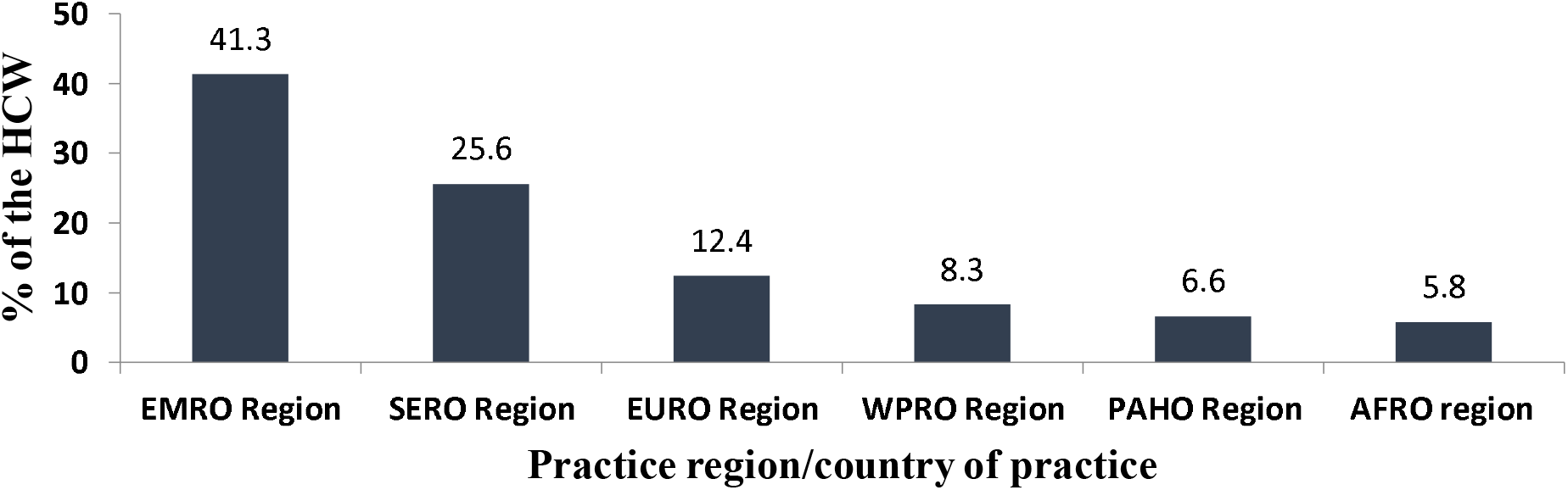
The distribution of the areas of practice of the responding healthcare workers.

### The nature of COVID-19 simulations or dills based on respondents

The nature and composition of COVID-19 simulations are shown in Table 2. The institutions varied in the intensity of drills conducted till the survey period (14 April 2020-27 May 2020), 28.9% of the respondents indicated that their hospitals had conducted more than 10 COVID-19 simulation sessions while 18% conducted only one session, and 37.4% conducted 2-4 sessions.

**Table 2:**
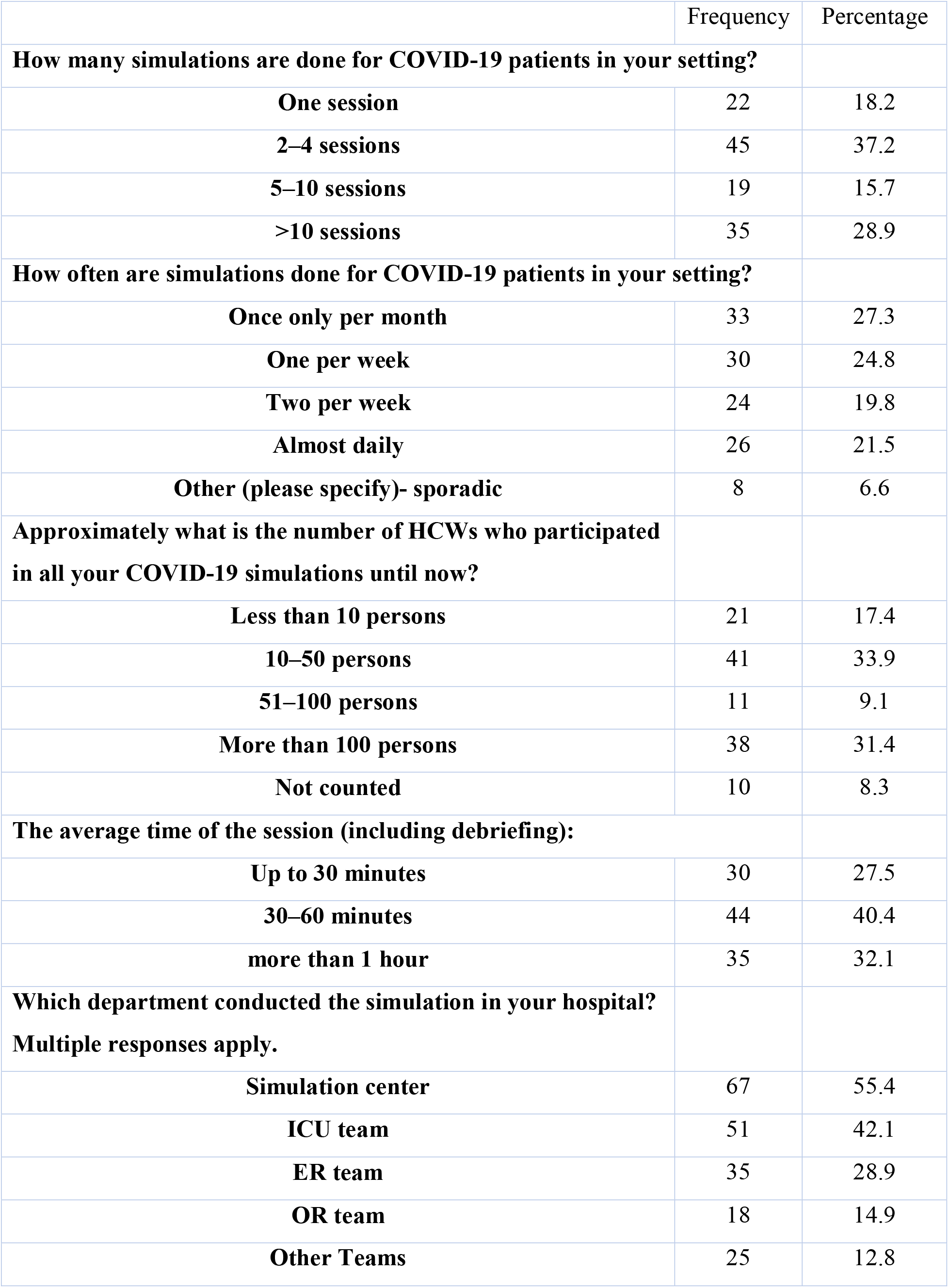
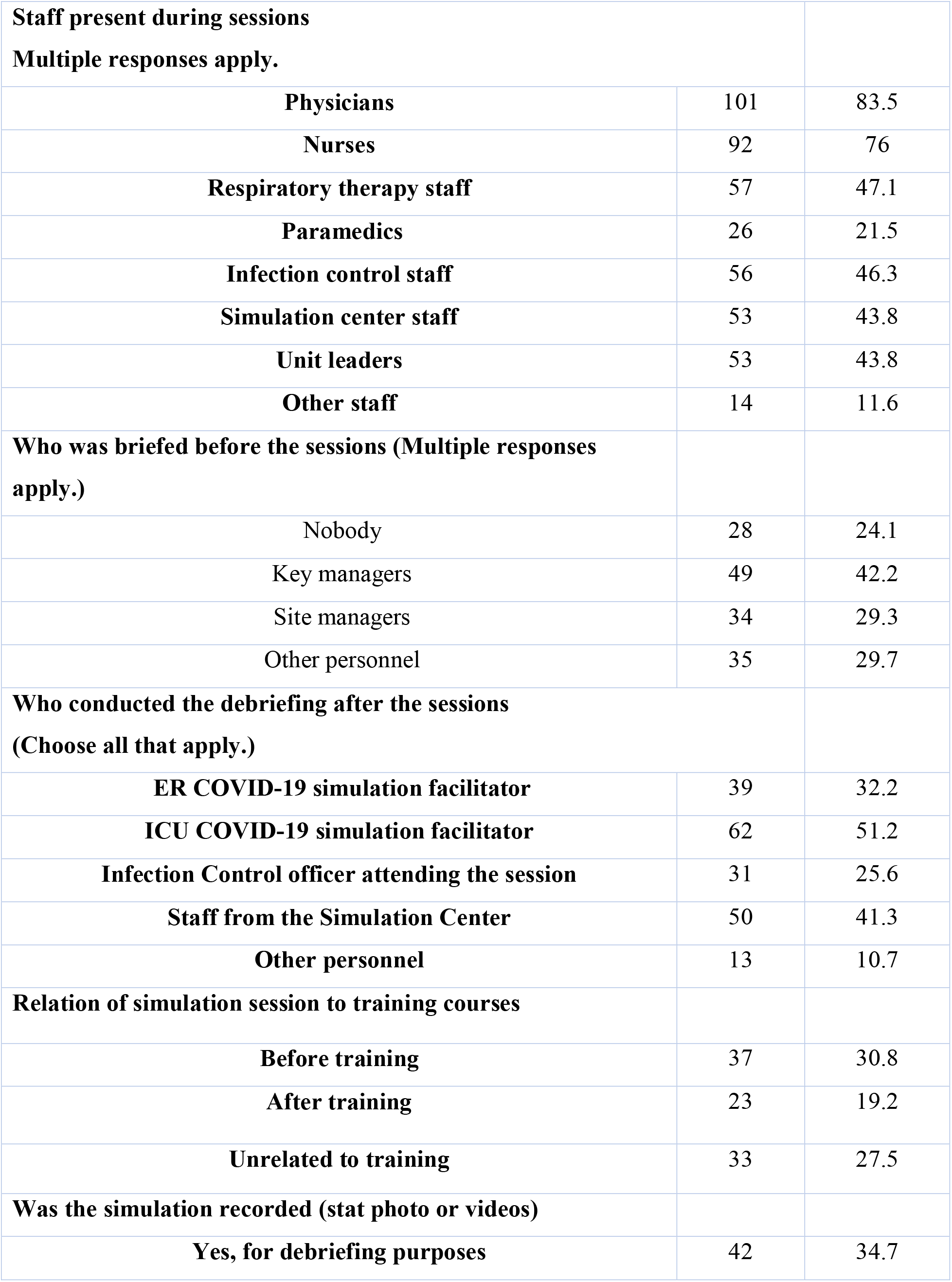
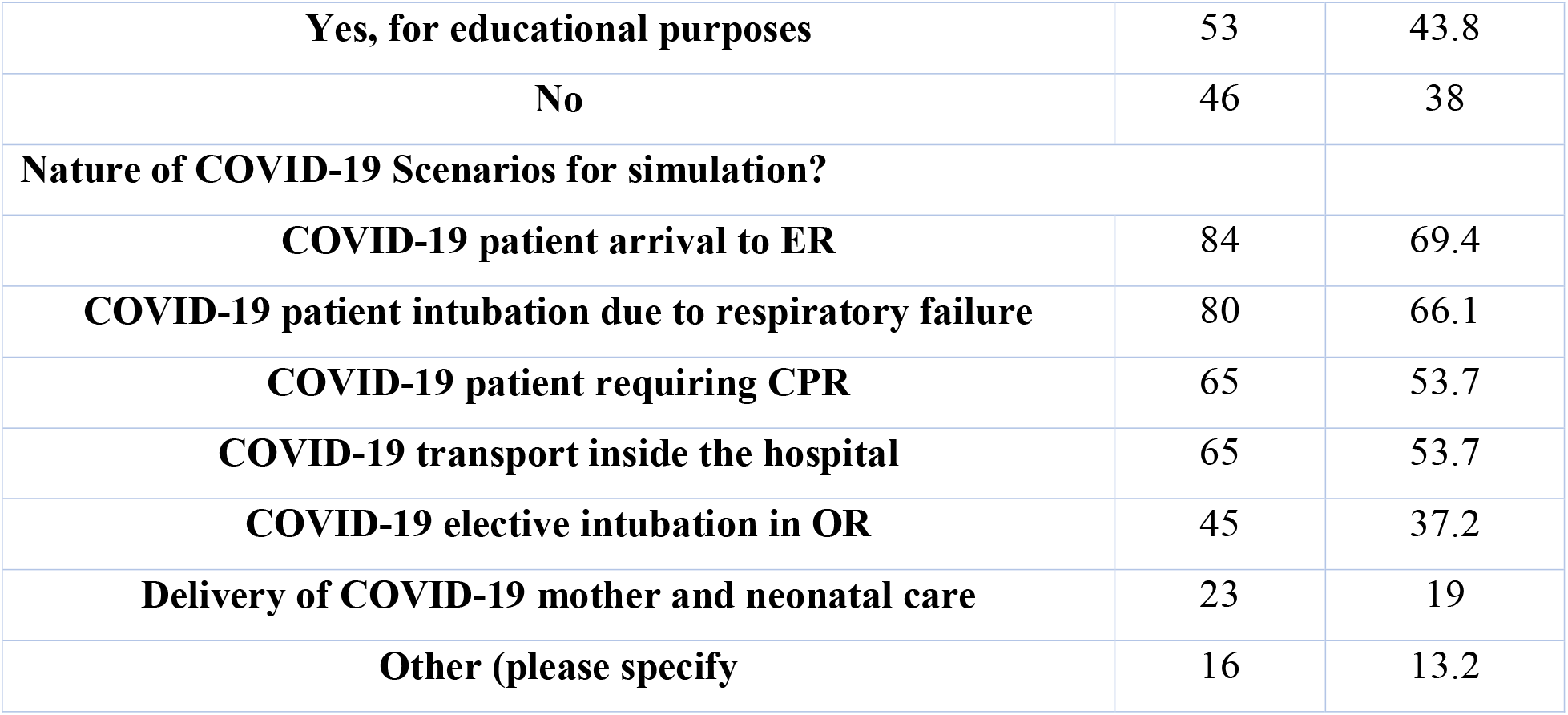
Description of COVID 19 simulation sessions (Drills) based on respondents. N = 121.

The frequency of the sessions was as follows: monthly (27.1%), weekly (24.8%), twice weekly (19.8%), and daily (21.5%). There was a significant variation in the number of personnel attending the sessions across the centers, one-third reported 10–50 people participated in the sessions, another third reported that more than 100 people participated in the COVID-19 simulation sessions in their hospitals, while 17.4% reported less than 10 personnel participating in the sessions.

The average length of each session was 30–60 minutes for 40% of respondents while for sessions less than 30 minutes or more than an hour each were conducted in one third of the centers. Most of the sessions were conducted in and by staff from the simulation center, followed by the ICU and emergency room (Figure 2).

**Figure 2:**
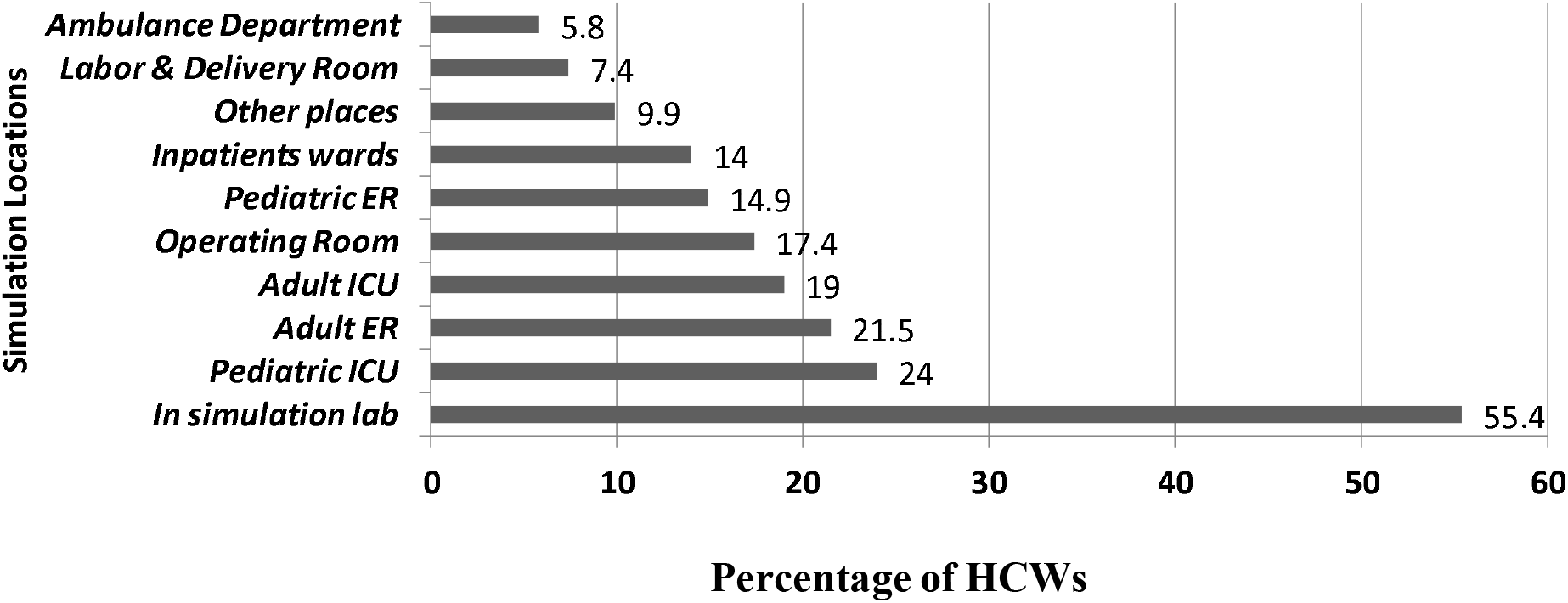
Locations where the COVID-19 drills were held, according to the healthcare workers.

The majority of sessions attendants were physicians and nurses followed by respiratory therapists and staff from infection control (Figure 3).

**Figure 3:**
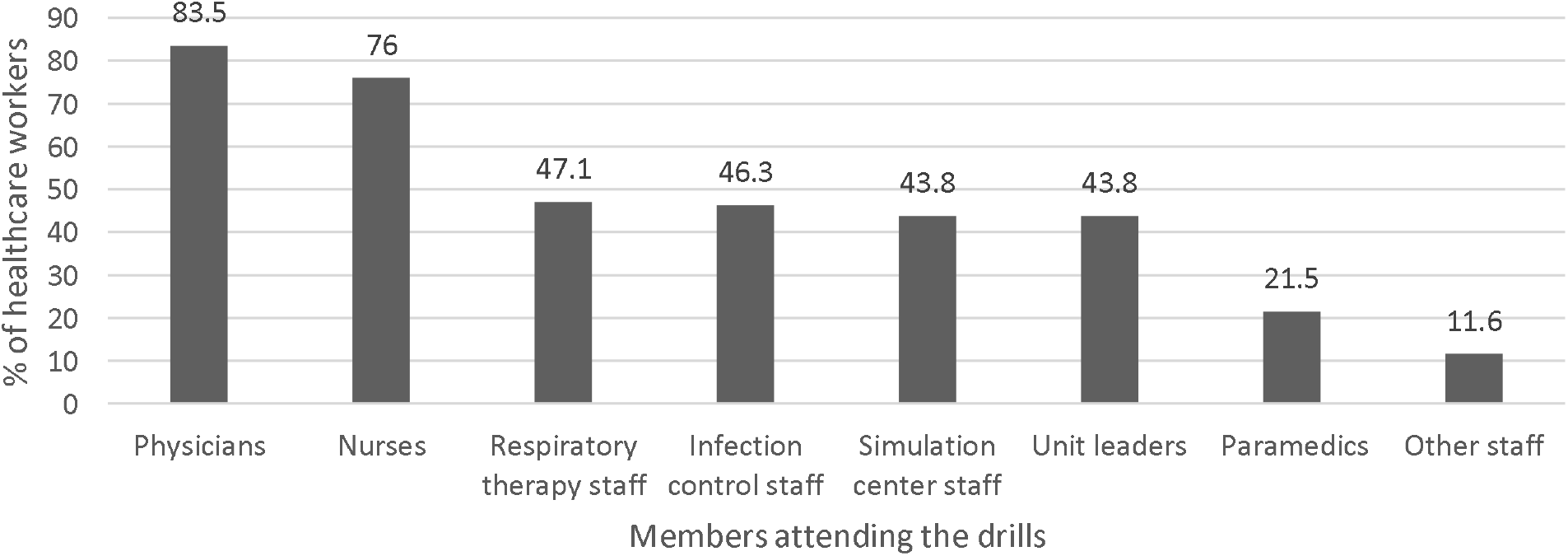
Composition of the drill teams according to the healthcare workers.

### Briefing and scenario selection

In 25% of the session no one was briefed before the session, while 42.2% reported that key managers were present, 29.3% advised that site managers were present, and 19% reported that other hospital personnel were involved in the preparatory briefing.

In relation to the debriefing process, it was mostly conducted by ICU facilitator (51%) followed simulation staff (41%) (Table 2).

The simulation sessions were mainly conducted before training courses in (30.8%) or after courses in (19.2%), while in 27.5% of the cases were unrelated to training courses. Most simulation sessions were recorded for debriefing or education purposes. Different scenarios (e.g., elective intubation, transport, etc.) were used, as detailed in Table 2.

### Respondents perceptions about their institutional COVID-19 simulations

Table 3 illustrates the respondents’ perceptions and attitudes about their institutions’ COVID-19 simulation sessions. Regarding simulation sessions improvement of institutional preparedness to COVID 19, 5% perceived minimal improvement, 16.5% perceived moderate improvement, 80% reported “a lot of” or “a great deal of” clinical preparedness improvement after the sessions.

**Table 3:** Healthcare workers’ perceptions and attitudes about their institutions’ COVID-19 simulation sessions. N = 121.

|  | Frequency | Percentage |
| --- | --- | --- |
| How much did COVID-19 simulation improve your own healthcare preparedness? |  |  |
| Minimal | 6 | 5 |
| A moderate amount | 20 | 16.5 |
| A lot | 47 | 38.8 |
| To a great deal | 48 | 39.7 |
| How engaged were the participating staff with the simulation |  |  |
| Minimal engagement | 3 | 2.4 |
| A little engaged | 1 | 0.8 |
| Moderately engaged | 39 | 31 |
| Highly engaged | 13 | 10.3 |
| Fully engaged | 70 | 55.6 |
| Overall, how would you rate the interactions of the simulation audience during the practice drill? |  |  |
| Fair | 2 | 1.7 |
| Good | 27 | 22.3 |
| Very Good | 56 | 46.3 |
| Excellent | 36 | 29.8 |
| How well did COVID-19 simulations meet your expectations? |  |  |
| Much worse than expected | 1 | 0.8 |
| Worse than expected | 1 | 0.8 |
| About the expected | 32 | 26.4 |
| Better than expected | 53 | 43.8 |
| Much better than expected | 34 | 28.1 |
| Do you think COVID-19 drill you conducted was too long, too short, or about right? |  |  |
| Much too short | 1 | 0.8 |
| Too short | 4 | 3.3 |
| About right length | 108 | 89.3 |
| Too Long | 8 | 6.6 |
| Did you face any risky encounter during your COVID-19 simulations? |  |  |
| Yes (please specify) | 14 | 11.6 |
| No | 107 | 88.4 |

Most of the HCWs (55.6%) reported full engagement by themselves and their peers with the sessions, another 31% reported moderate while only 10.3% high engagement. Almost half (43.8%) of respondents felt sessions were better or much better (28.1%) than expected. The majority of respondents felt session were of appropriate duration. Interestingly (11.6%) indicated that they encountered what they considered risky encounter during COVID-19 simulation session, such as no enough social distancing during the drill or insufficient surface sanitizing.

Figure 4 shows respondents responses regarding time required to put on PPE during COVID emergency simulation sessions, 9.2% required one minute or less, 23.9% 2 minutes, 22% 3 minutes, while the rest needed longer durations.

**Figure 4:**
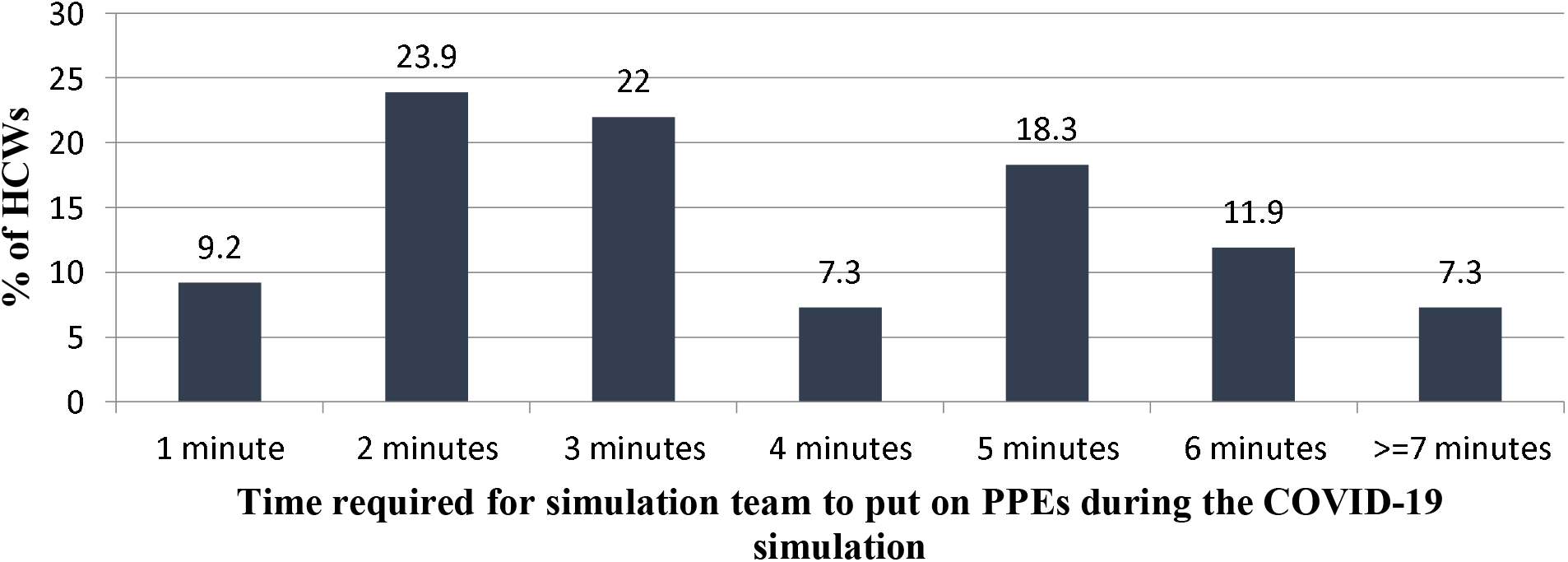
The distribution of time required for healthcare workers to put on personal protection equipment during COVID-19 simulations/drills.

### Problems perceived during COVID-19 simulations

The HCWs were asked to rate the frequency of challenges (issues) or problems encountered during the simulations of COVID-19 scenarios (Table 4). These issues fell into four categories:

**Table 4:**
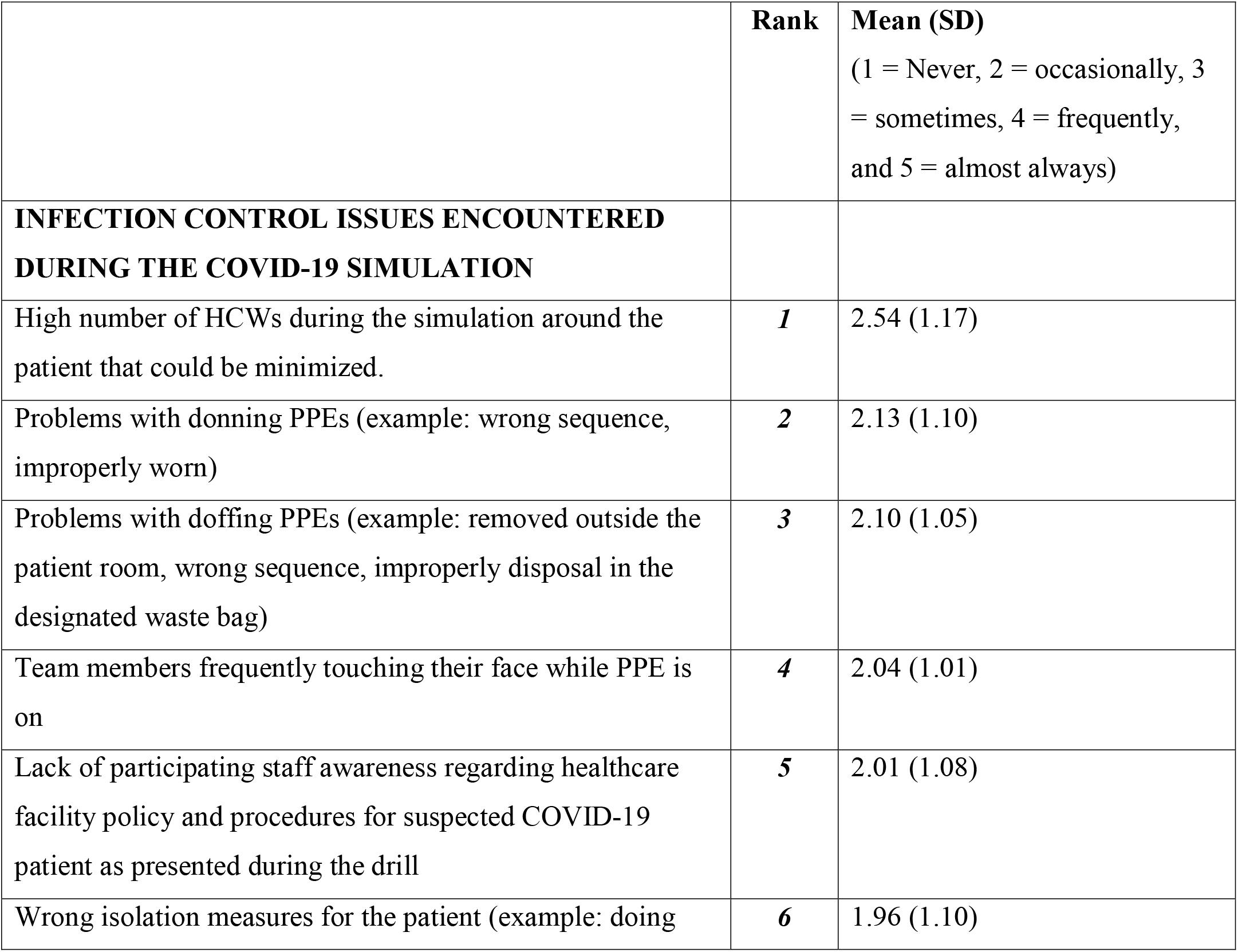
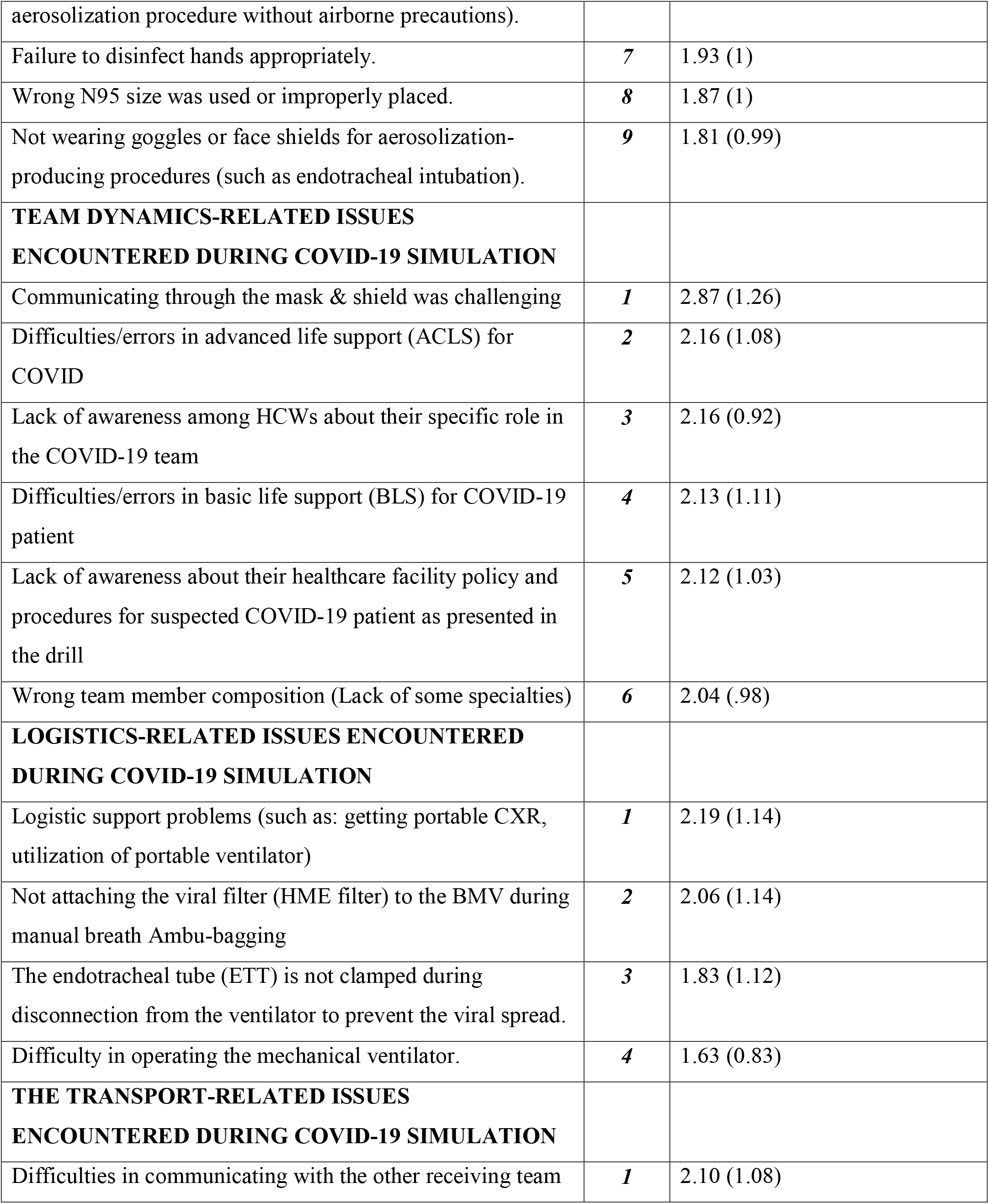
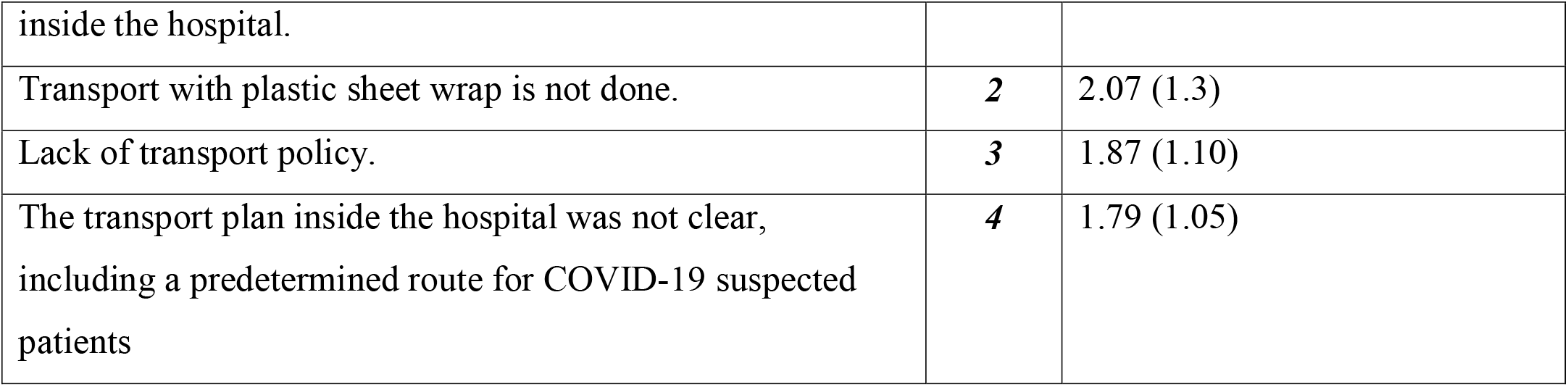
Descriptive analysis of issues and problems perceived during COVID-19 simulation.

A. Infection Control-Related Issues: The top infection control-related issue faced during the simulations was the high number of HCWs attending the patient during the simulation, followed by problems with donning the personal protective equipment (PPE) and doffing the PPE’s, and not attaching the HME viral filter during Ambu-bag ventilation, and difficulties related to the operation of the mechanical ventilators and “not clamping” the endotracheal tube during the disconnect period to prevent viral spread from the patients to the surrounding. Other issues are described in Table 4.
B. Team Dynamics-Related Issues: The top three perceived team dynamics issues encountered were communicating while wearing the face mask and face shields, followed by advanced life support errors and challenges when dealing with COVID-19 patients especially in relation to intubation and handling airways, and the lack of awareness among participants about their specific roles in the COVID-19 team.
C. Logistics-Related Issues: The top perceived issues associated with logistics during the COVID-19 simulations were logistics-related issues such as getting portable chest x-rays done and use of ventilators and related equipment.
D. Patient Transport-Related Issues: The top perceived COVID-19 transport-related issue was the difficulty in communicating with the receiving team in relation to the transport, issues with plastic sheet wrap-up being incomplete or not used at all; other issues are shown in Table 4.

### HCWs’ perceived challenges and potential facilitators for successful COVID-19 simulations

Respondents were asked to select from a list of challenges and difficulties that pressurized their simulation and facilitating factors that enhanced their debriefing. The resulting multiple response findings are displayed in Table 5.

**Table 5:** Respondents perceived challenges and potential facilitators for successful COVID- 19 simulations. N = 120.

|  | Frequency | Percentage |
| --- | --- | --- |
| **CHALLENGES FACED DURING THE COVID-19 SIMULATION** |  |  |
| Risk of crowded HCWs during the simulation session while social distancing is advised | 63 | 52.5 |
| Depletion of PPEs while there is a limited stock during the pandemic | 63 | 52.5 |
| A high number of hospital units asking for COVID-19 simulation drills beyond institution resources | 35 | 29.2 |
| Busy Infection Control colleagues unable to attend your drill | 27 | 22.5 |
| Busy simulation center staff unable to attend your drills | 23 | 19.2 |
| Lack of administrative support for COVID-19 simulation | 13 | 10.8 |
| Other challenges | 9 | 7.5 |
| **WHAT MADE YOUR SIMULATION/ DEBRIEFING OUTCOME EFFICIENT** |  |  |
| Scheduled drill so the HCWs could get ready | 64 | 53.3 |
| Both sudden or scheduled drills showed important findings for debriefing | 45 | 37.5 |
| Making the drill sudden without previous knowledge of the HCWs | 28 | 23.3 |

The most common challenge was non-compliance or difficulty to comply with social distancing during COVID 19 drills or real cases (52.5%) followed by limited resources to conduct drills across the different units of the institution (29.2%), followed by limitation of attendance of key personnel during the simulation like infection control officer or simulation trainer or administrative representative.. Other less frequently reported issues are listed in Table 5. We surveyed respondents regarding anxiety of PPE depletion during COVID pandemic and 52.5% reported their concern in that regard.

Regarding facilitator of successful simulation, 53.3% of the respondents felt that a scheduled drill is a facilitator of success especially staff had time to get ready. While 23.3% felt sudden drills are better facilitator for success, on the other hand 37.5% felt that drills either sudden or scheduled showed important finding for feedback and debriefing to improve future experience.

## Discussion

For more than 20 years, we have been witnessing a slow cultural change in healthcare; one that moves safe procedures into focus. While different methods are being used to enhance the safety of the whole system, thus directly influencing patient and HCW safety, simulation has been one of the key elements in this quest. The most common way of demonstrating and learning about safety culture is simulation training [(Motola et al., 2013)].

Although today no other high risk profession training exist without simulation, the progress in healthcare simulation has not been as rapid as initially predicted [(Motola et al., 2013)]. This might be due to number of factors, most prominently that while most other professions implementing a wide use of simulation are heavily based on technology, healthcare systems operate on humans which makes simulation challenging to apply [(Catchpole, 2013, Lazarovici et al., 2017)]. Simulation training is a valuable method for uncovering latent safety threats in healthcare systems [(Andreatta et al., 2011, Patterson et al., 2013, Wetzel et al., 2013)]. The use of simulation during the COVID-19 crisis could be particularly useful in this regard [(Aldekhyl and Arabi, 2020, Lababidi et al., 2020)].

Owing to its superior ability to detect latent safety threats, *in situ* simulation has been more widely used during recent years, while this development varies on a global level [(Patterson et al., 2013, Weinstock et al., 2009)]. Collaborative multicenter research offers many advantages over single-center simulation debriefing, including larger sample sizes for more generalizable findings, the ability to share findings amongst collaborative simulation sites, and networking capabilities [(Cheng et al., 2017)]. However, the response rates we achieved from various international regions might indicate that the numbers of COVID-19 simulations vary across various settings.

According to our study about 45% of the surveyed centers conducted at least 5 COVID-19 drills till May 2020 and in 40% of the cases it was conducted twice weekly or even daily since the announcement of the pandemic which reflects high alert level withing few months of the announcement across the healthcare system. Some simulation centers reported conducting more than 100 COVID-19 simulation scenarios, both *in situ* and in their simulation labs [(Cheung et al., 2020)]. When centralized regional COVID-19 simulations were implemented, the number of simulations was rapidly increased to more than 400 acute care simulation session requests across Alberta’s broad geographical zones within five□weeks [(Dubé et al., 2020)].

The sessions involved at least 50 personnel in total and reached more than 100 in 30% of the surveyed institutes, in at least 75% of the surveyed institutes the sessions lasted at least 30 minutes and more than an hour in about 10%, those facts reflect the effective and highly resourced planning of those sessions in many centers withing few months of the global alert.

Almost 90% of the drills were conducted either in simulation centers, ER or ICU setting and by their staff, pointing that those institutes took COVID 19 drills at high consideration level and conducted the drills at portal of entry of such patients or in areas where handling them might go hectic especially at the level of airways or infection control necessitating high level of preparedness which is reflected by the nature of the drills as detailed in Table 2.

The drills were well conducted most of the time as reflected by staff satisfaction and feeling of improvement post drills pointing the efficacy of those drills to face the pandemic and to reassure the anxious staff. Fear can hamper performance and may hinder learning. It is important that HCWs feel safe while learning, and a simulation scenario provides a safe environment to learn and practice a broad range of possibly hazardous situations [(Mohammadi et al., 2019)]. The feeling of fear regarding caring for infectious patients requiring strict isolation was reduced after the participants had completed the simulation and drill courses, as the majority of the HCW reported that their preparedness improved “a lot” to “a great deal” (78.5%) in comparison to those (5%) for whom preparedness improved only a little. Based on these findings, simulation- based learning positively imparted confidence, capability, and knowledge to HCWs. When they feel safe, they will be able to deliver better care to the patients and protect themselves from acquiring the infection. Varying results were reported in other studies regarding perceptions of readiness after simulation-based training. Prescott and Garside reported that all the participants in their study felt better prepared for the assigned tasks after a simulation-based training [(Prescott and Garside, 2009)]. However, another study found that fewer participants felt prepared for tasks for which they had received simulation-based training [(Feingold et al., 2004)]. Our study showed a noteworthy improvement in perceptions of preparedness among the HCWs after the training. In a COVID-19 simulation assessment, Cheung et al. found significant improvement in all domains of personal strengths among 1,415 hospital staff members [(Cheung et al., 2020)]. 65.9% of the surveyed staff in our study were highly or fully engaged during the sessions in comparison to 3.2% for whom engagement was little and minimal, Khan and Kiani found that the participants in their simulation courses believed from the start that their colleagues who did not attend the course were less prepared to handle COVID-19 patients [(Khan and Kiani, 2020)]. At the end of the course, this perception was strengthened. This implies that respondents perceived that the course had better prepare them for the challenge of caring for COVID-19 patients compared to their colleagues who had not trained with them.

Our findings indicate variability in the video recordings of the COIVD-19 drills. Such video recordings, either for the debriefing or educational purposes, were also applied to the COVID-19 simulation. Ahmed et al. reported using video recordings of the session that were then played before the subsequent COVID-19 simulation learning session as a pre-briefing [(Ahmed et al., 2020)]. Mistakes in performance could be systematically identified and discussed among the participants.

The most frequent and highly ranked challenges in terms of infection control and team dynamics-related issues faced during the COVID-19 simulations were reflective of reality clinical practice challenges. For example, similarities between the reported HCWs crowdedness around the patient and issues with lack of compliance with infection control practices during *in situ* simulations in this study have been reported as leading causes of HCW-related infection in many healthcare systems’ COVID-19 outbreaks [(Ran et al., 2020, Sørensen et al., 2017)]. The same findings were noted by Erich Hanel et al. in their surveys [(Hanel et al., 2020)].

The top-ranking challenging team dynamics issues were related to the difficulty in communicating while masks and face shields on, application of advanced cardiac life support, and lack of role clarity in the newly formed COVID-19 teams. In addition, there were challenges with providing basic life support and institutional COVID-19 policy and procedures. A recent nationwide Canadian study identified many similar findings and challenges across both urban and rural health care settings and addressed simulation-based education to achieve system-based learning [(Dubé et al., 2020)].

The reported challenges during *in situ* simulation provide a rich source for individual, team, and institutional gap analysis and work as a need assessment tool. These challenges serve as a stimulus for rapid cycle deliberate practice, resulting in increased preparedness. Such an approach has been successfully used in many healthcare systems to prepare hospitals or a particular section of healthcare facilities during the COVID-19 crisis worldwide [(Dieckmann et al., 2020, Wong et al., 2020)].

Previous studies have demonstrated the utility of *in situ* simulation to advance healthcare provider skills and aid in the development of protocols and procedures [(Chaplin et al., 2020, Goldshtein et al., 2020)]. However, the use of simulation under the constraints inflicted by a pandemic needs further studies. Indeed, the COVID-19 pandemic poses new challenges to the execution of *in situ* simulation. These challenges include limited time, personnel, and personal protective equipment (PPE), with changing guidelines on appropriate use. Although simulation lab capacities are overwhelmed internationally during this pandemic, only 29% of HCWs considered this to be a challenge. Given these legitimate concerns, it is essential that simulation continues to be used. Despite these difficult times and to overcome these challenges, formats of combined *in situ* and virtual video-based simulation might offer the most protected, safe environment. This combination model also allows simulation leaders to identify and modify site-specific latent safety threats, which are system-based threats to patient safety that were not previously recognized [(Patterson et al., 2013)]. Moreover, this method offers a means of rapid knowledge dissemination, using very few resources and saving time, PPE, and simulation equipment, while allowing for social distancing, eliminating geography as a limitation to education delivery, and allowing use by HCWs in preparation for *in situ* simulation training, as more than half of them want to be prepared [(Coyne et al., 2018, Patterson et al., 2013)]. Of note, a recent COVID-19 simulation study found no significant differences between *in situ* and lab-based simulations for all domains of personal strengths that were assessed among their candidates [(Cheung et al., 2020)].

### Study limitations

We included centers with simulation centers in order to survey institutions trained already to simulation drills, this limits our study results as this pandemic is global pandemic affecting any healthcare institution.

Our questionnaire was available online worldwide, and we got replies from many countries, representing different regions of the globe. Some regions were represented more heavily, while we got few replies from others. South America was not represented in our sample, and in Europe, we received data only from Spain, the U.K., Italy, Germany, and Russia. Given the diversity of European healthcare systems and the varying degrees of impact of the COVID-19 pandemic in different European countries, the data may not represent the full bandwidth of the reality in European healthcare. Another region with representation that might lead to selection bias is Africa—our responses here are from Egypt, Morocco, and South Africa. While these countries reflect a certain diversity of healthcare systems on the continent, we would not assume that our data represent the whole African reality either.

While this is among the first international studies to explore the effects of several factors on COVID-19 simulations across all WHO regions, the number of responding simulation centers was relatively low. However, the information shared from all centers and HCWs was abundant. This study is subject to the limitations of cross-sectional surveys, including sampling, response, and recall biases. While we attempted to reach out to as many simulation centers as possible on the international level, there were many logistical difficulties in getting replies from several places, probably reflecting the busy healthcare system during this COVID-19 crisis [(Alleva et al., 2020)]. While the aim of our study was to explore lessons learned from COVID-19 in international simulations, still, to tailor this more, we suggest conducting similar studies in the future at national levels. That would explore more unique factors for each country and healthcare system.

## Conclusion

Globally, healthcare workers reported positive feedback from the COVID-19 simulations conducted *in situ* or in simulation labs. The presence of infection control personnel in the multidisciplinary team during the drill is warranted to highlight any latent hazards that could be addressed in advance. *In situ* simulation provides a valuable tool to rehearse the safe dynamics of HCWs on the frontline of COVID-19.

## Data Availability

Data is available upon reasonable request.

## Contributorship Statement

Mohamad Hani Temsah planned and conducted the study, and is responsible for the overall content as guarantor. Abdulkarim Alrabiaah, Abdullah Al Huzaimi, Fahad Al-Sohime analyzed the data and wrote the results and discussion. Ayman Al-Eyadhy and Nurah Alamro contributions in the method and the manuscript drafting and finalization. Vaibhavi Upadhye surveyed simulation centers. Fadi Aljamaan, Khalid Alhasan and Yaseen M Arabi contributions in the manuscript drafting and finalization. Marc Lazarovici and Abdulaziz Boker contributions in the study design, data collection and manuscript drafting and finalization. Amr Jamal finalized and submitted the paper.

## b. Funding

The authors are grateful to the Deanship of Scientific Research, King Saud University for funding through Vice Deanship of Scientific Research Chairs. The sponsor had no influence on the study design or the reporting of the results.

## c. Competing interests

The authors declare no conflict of interest.

## d. Acknowledgments

The authors wish to thank all contributing healthcare workers and leaders of the participating simulation centers for their valuable input. We also thank Hodhodat.com for the statistical analysis. The authors thank the Deanship of Scientific Research and RSSU at King Saud University for their technical support.

## Notes

### Competing Interest Statement

The authors have declared no competing interest.

### Author Declarations

Approved by the institutional review board at King Saud University, Riyadh, Saudi Arabia.

## References

1. IBM Corp. Released 2012. IBM SPSS Statistics for Windows, Version 21.0. Armonk, NY: IBM Corp.

2. Abrahamson SD, Canzian S, Brunet F. Using simulation for training and to change protocol during the outbreak of severe acute respiratory syndrome. Crit Care 2006;10(1):R3.

3. Ahmed OM, Belkhair AOM, Ganaw AEA, ElKersh MM, Adiga J. Anaesthesia simulation training during coronavirus pandemic: an experience to share. BMJ Simulation and Technology Enhanced Learning 2020:bmjstel-2020-000643.

4. Aldekhyl S, Arabi Y. Simulation role in preparing for COVID-19. Annals of Thoracic Medicine 2020;15(3):134–7.

5. Alleva G, Arbia G, Falorsi PD, Zuliani A. A sample approach to the estimation of the critical parameters of the SARS-CoV-2 epidemics: an operational design with a focus on the Italian health system. arXiv preprint arXiv:200406068 2020.

6. Andreatta P, Saxton E, Thompson M, Annich G. Simulation-based mock codes significantly correlate with improved pediatric patient cardiopulmonary arrest survival rates. Pediatr Crit Care Med 2011;12(1):33–8.

7. Catchpole K. Spreading human factors expertise in healthcare: untangling the knots in people and systems. BMJ Qual Saf. 22. England; 2013. p. 793–7.

8. Chaplin T, Thoma B, Petrosoniak A, Caners K, McColl T, Forristal C, et al. Simulation-based research in emergency medicine in Canada: Priorities and perspectives. Canadian Journal of Emergency Medicine 2020;22(1):103–11.

9. Cheng A, Kessler D, Mackinnon R, Chang TP, Nadkarni VM, Hunt EA, et al. Conducting multicenter research in healthcare simulation: Lessons learned from the INSPIRE network. Advances in Simulation 2017;2(1):6.

10. Cheung VK-L, So EH-K, Ng GW-Y, So S-S, Hung JL-K, Chia N-H. Investigating effects of healthcare simulation on personal strengths and organizational impacts for healthcare workers during COVID-19 pandemic: A cross-sectional study. Integrative Medicine Research 2020;9(3):100476.

11. Coyne E, Frommolt V, Rands H, Kain V, Mitchell M. Simulation videos presented in a blended learning platform to improve Australian nursing students’ knowledge of family assessment. Nurse education today 2018;66:96–102.

12. Dieckmann P, Torgeirsen K, Qvindesland SA, Thomas L, Bushell V, Langli Ersdal H. The use of simulation to prepare and improve responses to infectious disease outbreaks like COVID-19: practical tips and resources from Norway, Denmark, and the UK. Adv Simul (Lond) 2020;5:3.

13. Dubé M, Kaba A, Cronin T, Barnes S, Fuselli T, Grant V. COVID-19 pandemic preparation: using simulation for systems-based learning to prepare the largest healthcare workforce and system in Canada. Adv Simul (Lond) 2020;5:22.

14. Feingold CE, Calaluce M, Kallen MA. Computerized patient model and simulated clinical experiences: evaluation with baccalaureate nursing students. J Nurs Educ 2004;43(4):156–63.

15. Goldshtein D, Krensky C, Doshi S, Perelman VS. In situ simulation and its effects on patient outcomes: a systematic review. BMJ Simulation and Technology Enhanced Learning 2020;6(1):3–9.

16. Hanel E, Bilic M, Hassall K, Hastings M, Jazuli F, Ha M, et al. Virtual application of in situ simulation during a pandemic. Canadian Journal of Emergency Medicine 2020:1–4.

17. Khan JA, Kiani MRB. Impact of multi-professional simulation-based training on perceptions of safety and preparedness among health workers caring for coronavirus disease 2019 patients in Pakistan. Journal of Educational Evaluation for Health Professions 2020;17.

18. King DR, Patel MB, Feinstein AJ, Earle SA, Topp RF, Proctor KG. Simulation training for a mass casualty incident: two-year experience at the Army Trauma Training Center. J Trauma 2006;61(4):943–8.

19. Lababidi HM, Alzoraigi U, Almarshed AA, AlHarbi W, AlAmar M, Arab AA, et al. Simulation-Based training programme and preparedness testing for COVID-19 using system integration methodology. BMJ Simulation and Technology Enhanced Learning 2020:bmjstel-2020-000626.

20. Lazarovici M, Trentzsch H, Prückner S. [Human factors in medicine]. Anaesthesist 2017;66(1):63–80.

21. Mohammadi G, Tourdeh M, Ebrahimian A. Effect of simulation-based training method on the psychological health promotion in operating room students during the educational internship. Journal of Education and Health Promotion 2019;8.

22. Motola I, Devine LA, Chung HS, Sullivan JE, Issenberg SB. Simulation in healthcare education: a best evidence practical guide. AMEE Guide No. 82. Med Teach 2013;35(10):e1511–30.

23. Organization WH. Rolling updates on coronavirus disease (COVID-19); 2020. Available from: https://www.who.int/emergencies/diseases/novel-coronavirus-2019/events-as-they-happen. [Accessed 2 July 2020.

24. Patterson MD, Geis GL, Falcone RA, LeMaster T, Wears RL. In situ simulation: detection of safety threats and teamwork training in a high risk emergency department. BMJ Qual Saf 2013;22(6):468–77.

25. Prescott S, Garside J. An evaluation of simulated clinical practice for adult branch students. Nursing Standard (through 2013) 2009;23(22):35.

26. Ran L, Chen X, Wang Y, Wu W, Zhang L, Tan X. Risk Factors of Healthcare Workers with Corona Virus Disease 2019: A Retrospective Cohort Study in a Designated Hospital of Wuhan in China. Clin Infect Dis 2020.

27. Richards GA, Sprung CL. Chapter 9. Educational process. Recommendations and standard operating procedures for intensive care unit and hospital preparations for an influenza epidemic or mass disaster. Intensive Care Med 2010;36 Suppl 1(Suppl 1):S70-9.

28. Sprung CL, Zimmerman JL, Christian MD, Joynt GM, Hick JL, Taylor B, et al. Recommendations for intensive care unit and hospital preparations for an influenza epidemic or mass disaster: summary report of the European Society of Intensive Care Medicine’s Task Force for intensive care unit triage during an influenza epidemic or mass disaster. Intensive Care Med 2010;36(3):428–43.

29. Sørensen JL, Østergaard D, LeBlanc V, Ottesen B, Konge L, Dieckmann P, et al. Design of simulation-based medical education and advantages and disadvantages of in situ simulation versus off-site simulation. BMC Med Educ. 17; 2017. p. 20.

30. Verbeek JH, Rajamaki B, Ijaz S, Tikka C, Ruotsalainen JH, Edmond MB, et al. Personal protective equipment for preventing highly infectious diseases due to exposure to contaminated body fluids in healthcare staff. Cochrane Database Syst Rev 2019;7(7):Cd011621.

31. Weinstock PH, Kappus LJ, Garden A, Burns JP. Simulation at the point of care: reduced-cost, in situ training via a mobile cart. Pediatr Crit Care Med 2009;10(2):176–81.

32. Wetzel EA, Lang TR, Pendergrass TL, Taylor RG, Geis GL. Identification of latent safety threats using high-fidelity simulation-based training with multidisciplinary neonatology teams. Jt Comm J Qual Patient Saf 2013;39(6):268–73.

33. WHO. Alphabetical List of WHO Member States; Available from: https://www.who.int/choice/demography/by_country/en/. [Accessed 20 October 2020.

34. Wong J, Goh QY, Tan Z, Lie SA, Tay YC, Ng SY, et al. Preparing for a COVID-19 pandemic: a review of operating room outbreak response measures in a large tertiary hospital in Singapore. Can J Anaesth 2020;67(6):732–45.

